# Male-Female Disparities in Years of Potential Life Lost Attributable to COVID-19 in the United States: A State-by-State Analysis

**DOI:** 10.1101/2021.05.02.21256495

**Authors:** Jay J. Xu, Jarvis T. Chen, Thomas R. Belin, Ronald S. Brookmeyer, Marc A. Suchard, Christina M. Ramirez

## Abstract

Males are at higher risk relative to females of severe outcomes following COVID-19 infection. Focusing on COVID-19-attributable mortality in the United States (U.S.), we quantify and contrast years of potential life lost (YPLL) attributable to COVID-19 by sex based on data from the U.S. National Center for Health Statistics as of 31 March 2021, specifically by contrasting male and female percentages of total YPLL with their respective percent population shares and calculating age-adjusted male-to-female YPLL rate ratios both nationally and for each of the 50 states and the District of Columbia. Using YPLL before age 75 to anchor comparisons between males and females and a novel Monte Carlo simulation procedure to perform estimation and uncertainty quantification, our results reveal a near-universal pattern across states of higher COVID-19-attributable YPLL among males compared to females. Furthermore, the disproportionately high COVID-19 mortality burden among males is generally more pronounced when measuring mortality in terms of YPLL compared to age-irrespective death counts, reflecting dual phenomena of males dying from COVID-19 at higher rates and at systematically younger ages relative to females. The U.S. COVID-19 epidemic also offers lessons underscoring the importance of a public health environment that recognizes sex-specific needs as well as different patterns in risk factors, health behaviors, and responses to interventions between men and women. Public health strategies incorporating focused efforts to increase COVID-19 vaccinations among men are particularly urged.

## 1. Introduction

Severe acute respiratory syndrome coronavirus 2 (SARS-CoV-2) [1], the beta-coronavirus that causes what is known as coronavirus disease 2019 (COVID-19), was first identified in an outbreak in Wuhan, Hubei province, China in December 2019. The virus rapidly spread throughout the world, and on 11 March 2020, the World Health Organization officially declared the international COVID-19 situation a pandemic [2]. In the United States (U.S.), the first confirmed case was identified on 20 January 2020 [3,4] in Washington state, although evidence suggests COVID-19 may have arrived in the U.S. as early as December 2019 [5,6]. The number of infected individuals and subsequent deaths quickly grew, and as of 31 March 2021—slightly past the one-year anniversary of the pandemic—the U.S. had over 30.4 million cumulative confirmed COVID-19 cases and 551,660 COVID-19 deaths [7], both figures the highest among every country in the world [8].

Clinical studies on COVID-19 patients conducted early in the pandemic found that men were dying at markedly higher rates relative to women [9–13]. At the population level, males comprise the majority of COVID-19 deaths in the overwhelming majority of countries that report sex-disaggregated COVID-19 mortality data [14]. Here, we focus on the U.S., where the majority of COVID-19 deaths also occur among males [15]. Standard analyses and commentaries contrasting the male and female population level COVID-19 mortality burdens typically involve calculating the percentage of total deaths by sex—contrasting them with their respective percent population shares—and/or calculating male and female (age-adjusted) mortality rates [16–18]. For example, the GenderSci Lab COVID Project at Harvard University [18] tracks the number of male and female COVID-19 deaths by state, calculating the percentage of total deaths by sex as well as crude and age-adjusted male and female mortality rates for each state. However, because COVID-19 case fatality rates are considerably higher among individuals in older age groups, COVID-19 death counts and mortality rates for both males and females are predominantly determined by data from COVID-19 decedents in older age groups. Younger individuals, however, are also susceptible to death from COVID-19, which in principle represent greater unrealized years of life, economic productivity, and broader contributions to society compared to decedents of greater age.

Years of potential life lost (YPLL) is a widely used epidemiological measure of mortality burden that emphasizes deaths that occur at younger ages by explicitly weighting such deaths more heavily [19]. The mathematical formula for YPLL for an individual fatality *i* is defined to be the difference between an upper reference age 𝒜 (typically close to a widely applicable life expectancy) and age at death *a*_*i*_ if the difference is positive and zero otherwise:

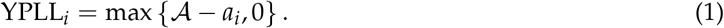

Here, we quantify disparities in YPLL attributable to COVID-19 in the U.S. by sex at the state level to examine both their magnitudes and their state-to-state variation. Specifically, we characterize the disparities by estimating (a) percentages of total YPLL by sex and (b) age-adjusted male-to-female YPLL rate ratios (RR), both nationally and for each of the 50 states and the District of Columbia (D.C.). For comparison, we also calculate the corresponding percentages of total deaths by sex and estimate the corresponding age-adjusted male-to-female mortality RR’s to examine potential differences in the characterization of the disparities when measuring mortality in terms of YPLL compared to (age-irrespective) death counts. To perform estimation and uncertainty quantification of the quantities of interest, we use novel Monte Carlo (MC) simulation techniques to obtain interval estimates for them.

## 2. Materials and Methods

### 2.1 Data

We examine U.S. national COVID-19 mortality data from the National Center for Health Statistics (NCHS) summarized as cumulative death counts within age intervals stratified by state (as well as D.C. and Puerto Rico) and sex [20]. The sex categories are male and female, and the following set of mutually exclusive, collectively exhaustive, and chronologically ordered age groups are used: <1, 1–4, 5–14, 15–24, 25–34, 35–44, 45–54, 55–64, 65–74, 75–84 and 85+. Death counts between 1 and 9 are suppressed in the NCHS data due to patient privacy laws. However, the NCHS data additionally provides the total number of male and female deaths in each jurisdiction. Therefore, for each state and sex, we know the total number of deaths that are within the union of age groups with suppressed death counts, each of which contains between 1 and 9 deaths. The NCHS data also provides non-suppressed death counts within these same age groups stratified by sex for the U.S. overall.

See File S1 in the Supplementary Materials for the NCHS data as of 31 March 2021 (reflecting all COVID-19 deaths reported to the NCHS as of 27 March 2021), which comprises 533,291 total deaths and represents 97.2% of the actual number of COVID-19 deaths—548,403 according to the New York Times [7]—in the U.S. as of 27 March 2021. The difference arises because there is a lag in time between the actual date of death and when the death certificate is completed, submitted to NCHS, and processed. A total of 111 male age groups and 113 female age groups have suppressed death counts across the 50 states and the District of Columbia (D.C.). COVID-19 deaths in a small number of states have been documented as being reported to the NCHS in a severely delayed timeframe [21–23]. North Carolina in particular is severely delayed in reporting COVID-19 deaths to the NCHS because, at the time of writing, it is one of a few states that still does not use an electronic death registration system [24]. As of 27 March 2021, North Carolina has had 12,049 COVID-19 deaths [7], but the NCHS data as of the same date contains only 6,378 (52.9%) deaths in North Carolina. Regardless of the degree of lag in COVID-19 death reporting to the NCHS in each state, our analysis implicitly assumes in each state that there is no systematic bias in the speed with which death certificates are reported by states to the NCHS with respect to sex or age at death.

To standardize estimates by age, we use the 2019 U.S. Census Bureau estimates of the population age distribution in each state (and D.C.) by sex [25], defined over integer ages from 0 to 84 and a catch-all 85+ age group for the remaining ages. See File S2 in the Supplementary Materials for the 2019 U.S. Census Bureau data.

### 2.2. Previous Work Quantifying Male-Female Disparities in COVID-19-Attributable YPLL in the United States

YPLL has been used in diverse contexts to quantify and contrast the impact of premature mortality by sex (e.g., [26–35]), and it has been used as a quantitative measure of mortality burden in the context of COVID-19 (e.g., [36–44]). In particular, Quast et al. (2021) [45] analyzed NCHS data as of 3 February 2021 (reflecting all COVID-19 deaths reported to the NCHS as of 31 January 2021)—serving as a rough approximation of U.S. COVID-19 deaths during the first year of the pandemic—estimating total YPLL and crude YPLL rates by sex in each U.S. state using the sex-specific remaining life expectancy method [46] to define YPLL, meaning that YPLL for each decedent is defined to be the expected number of remaining years of life conditional on sex and survival to the observed age at death with respect to the overall U.S. population. While their paper serves as a useful initial analysis contrasting COVID-19-attributable YPLL by sex, there are a number of methodological shortcomings in their study that we aim to address in our analysis.

First, they did not quantify the uncertainty of their state-level estimates of total YPLL and crude YPLL rates due to the administrative interval censoring of ages at death in the NCHS data. Quast et al. focused exclusively on point estimation, assuming for purposes of calculation that ages at death among decedents in a given age group all occurred at a fixed age, usually the age group midpoint (i.e., assuming that decedents in age group 60-69 died at age 65). For the 85+ age group in particular, because the ages at deaths are right-censored, they assumed all of these deaths occurred at age 90, an arbitrary assumption made purely out of analytical convenience to compute YPLL for these decedents. Second, they handled the suppressed death counts in the NCHS data by simply excluding them from their analysis, analogous to a “complete-case analysis” in the missing data literature [47], a suboptimal treatment of missing values that is susceptible to bias. Third, they did not account for differences in the male and female population age distributions within and between states by age-standardizing their YPLL rate estimates, especially important when a consistent comparison of the magnitudes of male-female disparities across states is desired. In Section 4, we also include a brief discussion of the methodological weakness of using the sex-specific remaining life expectancy method to define YPLL in the context of COVID-19. YPLL has been used to contrast male and female COVID-19-attributable YPLL in the state of Ohio [48], but outside of Quast et al. and to the best of our knowledge at the time of writing, YPLL has not yet been formally used as an epidemiological measure of mortality burden in the peer-reviewed literature to comprehensively characterize state-level disparities in the COVID-19 mortality burden between males and females in the U.S.

Our analysis improves upon the methodology employed in the Quast et al. study in three main ways. First, we define YPLL in a way that is practically useful, precedented, and compatible with the age groups used in the NCHS data. Second, we develop a novel adaptation of the MC simulation procedure proposed by Xu et al. (2021a) [49] to account for and quantify the estimation uncertainty arising from the administrative interval censoring of ages at death and the suppression of low death counts within individual age groups. And third, we account for differences in the male and female population age distributions within and between states by standardizing our male and female YPLL rate estimates to 2019 Census Bureau estimates of the U.S. national age distribution when estimating age-adjusted male-to-female YPLL RR’s both nationally and in each state (and D.C.).

### 2.3. Estimation Procedure for YPLL-Based Estimands from Administratively Interval Censored Ages at Death

As previously described in Section 1, we characterize disparities in COVID-19-attributable YPLL by sex through the estimation of percentages of total YPLL by sex—contrasting them with their respective percent population shares—and age-adjusted male-to-female YPLL RR’s. Additionally, to provide context into the magnitude of COVID-19-attributable YPLL experienced by males and females, we also perform estimation of total YPLL and age-adjusted YPLL rates by sex. As explained in Section 2.2, estimation uncertainty pertaining to the above YPLL-based estimands of interest can be attributed to two sources: (a) administrative interval censoring of ages at death and (b) suppression of low death counts within individual age groups.

We first focus on the issue of administrative interval censoring of ages at death, assuming momentarily there are no suppressed death counts for purposes of illustration. Because the exact ages at death for each individual are unknown, exact YPLL values for each individual are also unknown. In such settings, the standard approach to calculate aggregate YPLL is to operationally assume the age at death for each individual in a given age group is equal to the midpoint, also referred to as the “midpoint method,” which implicitly assumes that ages at death within each age group are uniformly distributed [50]. However, applied epidemiological studies using the midpoint method to estimate YPLL-based quantities typically do not quantify the uncertainty attributable to the administrative interval censoring of ages at death (e.g., [51–56]).

Xu et al. (2021a) [49] proposed a MC simulation procedure to quantify the uncertainty associated with YPLL-based estimates obtained from mortality data summarized as death counts within age intervals, which has been used in other applied research [44]. The full details of the procedure can be found in their paper, but to summarize it briefly, the Xu et al. MC simulation procedure consists of stochastic simulation of ages at death for each individual in the data from continuous uniform distributions defined over their respective age intervals at each MC iteration. A point estimate of the YPLL-based estimand of interest is then calculated from the collection of simulated ages at death at each MC iteration, and the overall point estimate is taken to be the mean of the collection of MC point estimates, while the lower and upper endpoints of a (1 − *α*) *×* 100% interval estimate (which can be conceptualized as a “range interval” per Bobashev and Morris (2010) [57]) are defined to be the 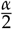 and 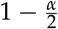 quantiles of the collection of MC point estimates, respectively.

### 2.4. Procedure Modification to Account for Suppression of Low Death Counts

We now address the second source of estimation uncertainty—suppression of low death counts within individual age groups—through a modification of the Xu et al. MC simulation procedure described in Section 2.3. A similar modification of the standard Xu et al. MC simulation procedure was also described in Xu et al. (2021b) [44] with respect to their comparative analysis of YPLL by race/ethnicity; as such, we use similar language in our description here. For each state and sex, we know the total number of deaths contained in the union of age groups with suppressed death counts, each of which must be an integer between 1 and 9. Hence, for each sex, we can exhaustively enumerate all possible death count combinations across the age groups with suppressed death counts. For each sex, each death count combination corresponding to the age groups with suppressed death counts juxtaposed with the age groups containing non-suppressed death counts constitutes one possible sex-specific “mortality dataset” of death counts within age groups.

We modify the Xu et al. MC simulation procedure described in Section 2.3 by independently simulating ages at death for each individual for each possible male-female mortality dataset pair at each MC iteration. Then, at each MC iteration, a point estimate of the estimand of interest is calculated from the simulated ages at death for each male-female mortlity dataset pair, from which we store only the minimum and maximum point estimates. A conservative (1 − *α*) *×* 100% interval estimate of the estimand of interest is then constructed from the 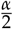 quantile of the collection of minimum MC point estimates and the 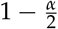 quantile of the collection of maximum MC point estimates. We describe the interval estimate as “conservative” due to our estimation strategy of enumerating all possible male-female mortality dataset pairs in the data and using the extrema of the subsequent MC point estimates to form an interval estimate. Indeed, if the suppressed death counts had actually been known, a (1 − *α*) *×* 100% interval estimate of the estimand of interest obtained from the standard Xu et al. MC simulation procedure [49] would be completely contained in the corresponding conservative (1 − *α*) *×* 100% interval estimate.

### 2.5. Computational Savings by Omitting Unnecessary Mortality Datasets

For each state and estimand of interest, the modified Xu et al. MC simulation procedure described in Section 2.4—in theory—comprises simulating ages at death for each individual in *B × J*_*ms*_ *× J*_*fs*_ male-female mortality dataset pairs, where *B* denotes the number of specified MC iterations, *J*_*ms*_ denotes the number of male mortality datasets in state *s*, and *J*_*fs*_ denotes the number of female mortality datasets in state *s*. As such, the total number of male-female mortality dataset pairs to simulate from can be enormous for sufficiently large values of either *J*_*ms*_ or *J*_*fs*_ (or both), potentially making it an excessively computationally burdensome endeavor. However, substantial computational savings can be achieved by identifying male-female mortality dataset pairs that we do not need to simulate ages at death from because they would yield a maximum or minimum MC point estimate of the estimand of interest with probability 0.

We describe an example of identifying such superfluous male-female mortality dataset pairs we can omit from our analysis when the estimand of interest is the male percentage of total YPLL. This is achieved by separately considering which male-female mortality dataset pairs will yield a maximum or minimum point estimate. To obtain a maximum point estimate at each MC iteration, only one male-female mortality dataset pair needs to be considered. The only male mortality dataset that needs to be considered is the one that contains the maximum possible number of deaths in the youngest age groups corresponding to suppressed death counts, and the only female mortality dataset that needs to be considered is the one that contains the maximum possible number of deaths in the oldest age groups corresponding to suppressed death counts. Any other male-female mortality dataset pair would yield a maximum MC point estimate with probability 0. A similar result applies when obtaining a minimum point estimate at each MC iteration. For that task, the only male mortality dataset that needs to be considered is the one that contains the maximum possible number of deaths in the oldest age groups corresponding to suppressed death counts, and the only female mortality dataset that needs to be considered is the one that contains the maximum possible number of deaths in the youngest age groups corresponding to suppressed death counts. Any other male-female mortality dataset pair would yield a minimum MC point estimate with probability 0. Hence, when the estimand of interest is the male percentage of total YPLL, only 2 male-female mortality dataset pairs need to be simulated from at each MC iteration.

For estimation of the age-adjusted YPLL-based estimands of interest (i.e., age-adjusted male and female YPLL rate, age-adjusted male-to-female YPLL RR), identifying male-female mortality dataset pairs that can be omitted is less straightforward. However, inequality conditions can be established computationally to identify meaningful numbers of male-female mortality dataset pairs that can validly be omitted, thereby attaining computational savings.

### 2.6. Complete Monte Carlo Simulation Procedure

Here, we comprehensively summarize the modified Xu et al. MC simulation procedure that we use in our analysis to perform estimation and uncertainty quantification of the YPLL-based estimands of interest. In summaries of the results of our analysis, we characterize D.C. as a “state” for brevity. For each state *s*, the procedure can be comprehensively summarized as follows.

1. Calculate the difference between the total number of male deaths and the number of male deaths contained in age groups with non-suppressed death counts. This difference is the number of male deaths contained in the union of age groups with suppressed death counts. Construct all possible male mortality datasets, each of which corresponds to a possible male death count combination across the age groups with suppressed death counts. Do the same for female deaths to obtain all possible female mortality datasets.

Let ℬ denote the total number of MC iterations, and let *b* = 1, …, ℬ index the MC iterations. Let *n*_*ms*_ denote the total number of male deaths in state *s*, and let *i*_*m*_ = 1, …, *n*_*ms*_ index the individual male deaths. Similarly, let *n*_*fs*_ denote the total number of female deaths in state *s*, and let *i* _*f*_ = 1, …, *n*_*fs*_ index the individual female deaths. Let *J ≤ J*_*ms*_ *× J* _*f s*_ denote the number of male-female mortality dataset pairs for state *s* that remain after omitting those male-female mortality dataset pairs that would yield a maximum or minimum point estimate with probability 0, and let *j* = 1, …, *J* index both their male and female mortality dataset constituents.

2. Specify a YPLL upper reference age *A* less than or equal to 85 years. We view age group <1 as equivalent to the singular age 0, and the remaining numeric NCHS age group endpoints represent integer age at last birthday so that there is a 1-year gap between the endpoints of two chronologically consecutive age groups (e.g., 35-44 and 45-54). We treat age as a continuous variable, and as a consequence, we mathematically interpret the <1 age group (age 0) as the right half-open interval [0, 1), the 85+ age group as the half-bounded interval [85, ∞), and the remaining NCHS age groups as right half-open intervals with lower limit equal to the lower endpoint of the corresponding NCHS age group and upper limit equal to the upper endpoint of the corresponding NCHS age group plus one (e.g., age group 15–24 is viewed as [15, 25)). Observe that *A* is intentionally and necessarily chosen to be less than or equal to 85 years to obviate the simulation of ages at death for decedents in the 85+ age group because each fatality in that age group contributes zero YPLL.

3. At each MC iteration *b*, independently simulate an age at death for each male decedent *i*_*m*_ in male mortality dataset *j* with reported age at death 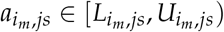 from a continuous uniform distribution over the same interval:

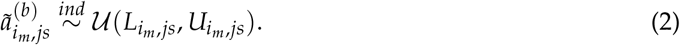

Likewise, independently simulate an age at death for each female *i* _*f*_ in female mortality dataset *j* with reported age at death 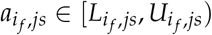 from a continuous uniform distribution over the same interval:

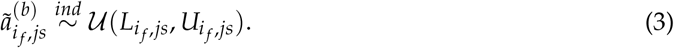

4. At each MC iteration *b*, calculate a point estimate of the estimand of interest from the simulated ages of death corresponding to each of the *J* male-female mortality dataset pairs. Specifically, for the estimation of the male percentage of total YPLL, first calculate total YPLL for males and females from the simulated ages at death, which are 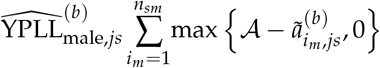 and 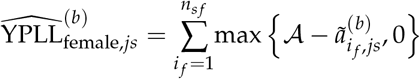, respectively. Then, the male percentage of total YPLL, which we denote 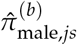, is given by:

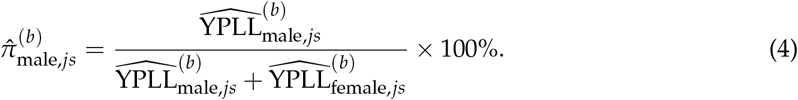

Similarly, the female percentage of total YPLL, which we denote 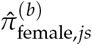, is given by

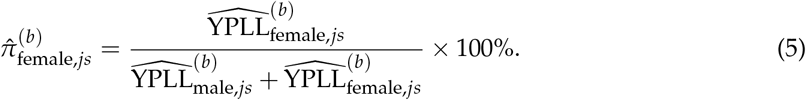

For estimation of the age-adjusted male-to-female YPLL RR, first estimate the age-adjusted male and female YPLL rates using direct age adjustment [58], using the 2019 U.S. Census Bureau age distribution estimate of the overall U.S. population as the standard population. Since the simulated ages at death are continuous and the U.S. Census Bureau age distribution estimates are defined over integer ages from 0 to 84, we aggregate the corresponding simulated YPLL values with respect to the 1-year age intervals implied by these integer ages (i.e., age *a* ∈ {0, 1, …, 84}implies age interval [*a, a* + 1)) to calculate the age-specific YPLL rates, which are subsequently applied to the standard population to obtain the age-adjusted YPLL rate. The male age-adjusted YPLL rate is given by:

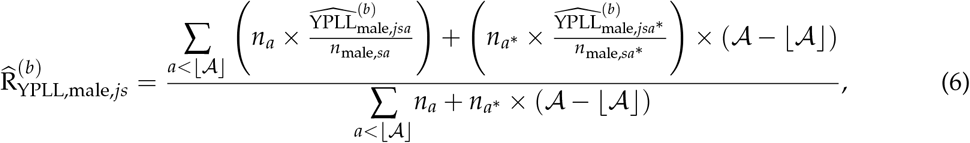

Where 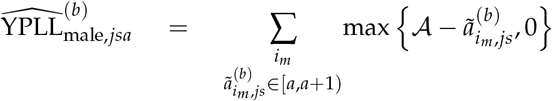, 0 denotes aggregate male YPLL corresponding to age *a* ∈ {0, 1, …, 84}; *n*_male,*sa*_ denotes the 2019 U.S. Census Bureau male population estimate for age *a* ∈ {0, 1, …, 84}in state *s*; *n*_*a*_ denotes the 2019 U.S. Census Bureau national population estimate for age *a* ∈ {0, 1, …, 84}; and *a*^∗^ = min{84, ⌊𝒜⌋}.

Analogously, the female age-adjusted YPLL rate is given by:

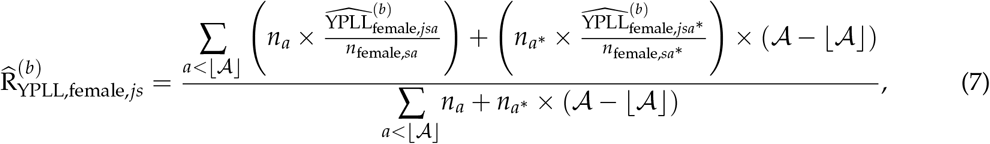

Where 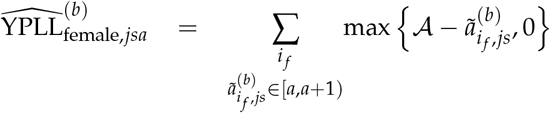 denotes aggregate female YPLL corresponding to age *a* ∈ {0, 1, …, 84}; and *n*_female,*sa*_ denotes the 2019 U.S. Census Bureau female population estimate for age *a* ∈ {0, 1, …, 84}in state *s*.

Then, the age-adjusted male-to-female YPLL RR is defined to be the quotient of 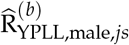 and 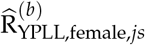

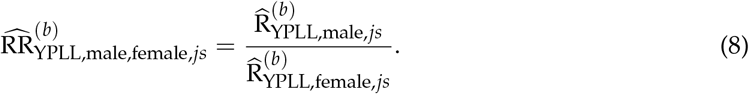

5. At each MC iteration *b*, store the maximum and minimum of the *J* MC point estimates of the estimand of interest obtained from the simulated male-female mortality dataset pairs.

6. A conservative (1 − *α*) *×* 100% interval estimate of the estimand of interest is given by the 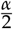 quantile of the ℬ minimum MC point estimates and the 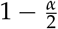 quantile of the ℬ maximum MC point estimates.

An overall point estimate for the estimand of interest is not straightforward to define as a result of our estimation strategy. To be explicit, the midpoint of the conservative (1 − *α*) *×* 100% interval estimate should not be interpreted as the point estimate. As such, we present the results of our analysis in terms of the collection of interval estimates we generate for the YPLL-based estimands of interest.

### 2.7. Monte Carlo Simulation Procedure for Estimation of Age-Adjusted Mortality Rates and Rate Ratios

We also consider estimation of age-adjusted mortality rates by sex and age-adjusted male-to-female mortality RR’s in the U.S. and in each state for the purpose of comparing them to our estimates of age-adjusted YPLL rates by sex and age-adjusted male-to-female YPLL RR’s, respectively, to examine potential differences in the characterization of the disparities when measuring mortality in terms of YPLL compared to (age-irrespective) death counts. Adopting the same methodological motivation as Xu et al. (2021b) in their estimation of age-adjusted mortality rates by race/ethnicity and their associated RR’s, we want our male and female mortality rates to be standardized to the 2019 U.S. Census age distribution estimate of the overall U.S. population—without combining age intervals in the U.S. Census data to align them with the NCHS data age intervals—so that estimated mortality and YPLL rates in our analysis are age-standardized to as identical as possible standard populations in terms of age interval granularity. To this end, we perform a MC simulation procedure to obtain conservative (1 − *α*) *×* 100% interval estimates of male and female age-adjusted mortality rates and age-adjusted male-to-female mortality RR’s that largely mirrors the MC simulation procedure for the YPLL-based estimands of interest described in Section 2.6. The key differences are that ages at death are simulated for all individuals in non-85+ age groups, and in the direct age adjustment procedure, we sum the number of simulated ages at death falling within the 1-year intervals implied by integer ages 0 to 84 to calculate the age-specific mortality rates for ages 0 to 84, as well as calculate an age 85+ mortality rate, which are subsequently applied to the standard population to obtain the male and female age-adjusted mortality rates, which we denote 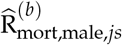 and 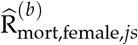 respectively.

Mathematically, the male age-adjusted mortality rate is given by:

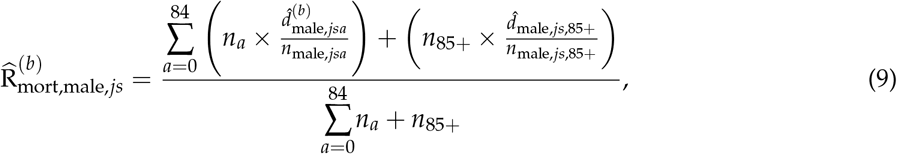

where 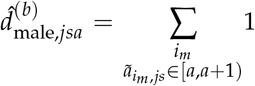 1 is the number of male simulated ages at death equal to age *a* ∈ {0, 1, …, 84}, 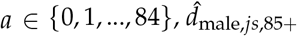 denotes the number of male deaths in the 85+ age group, and *n*_85+_ denotes the 2019 U.S. Census Bureau national population estimate for age group 85+.

Similarly, the female age-adjusted mortality rate is given by:

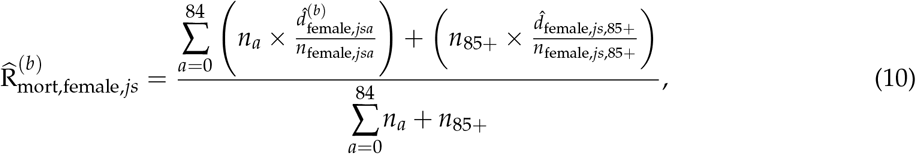

where 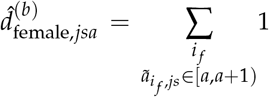 1 is the number of female simulated ages at death equal to age *a* ∈ {0, 1, …, 84}, and 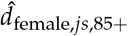 denotes the number of female deaths in the 85+ age group.

Then, the age-adjusted male-to-female mortality RR is defined to be the quotient of 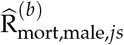 and 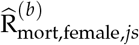

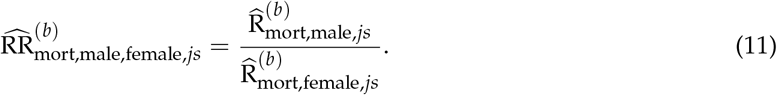

### 2.8. Computation

We perform the modified Xu et al. MC simulation procedure described in Section 2.6 for interval estimation of the YPLL-based estimands of interest in the U.S. and in each state. We perform the MC simulation procedure using ℬ = 1000 iterations and a constant YPLL upper reference age of 𝒜 = 75 years for both males and females, an approach used by the Centers for Disease Control and Prevention (CDC) [59–66] and widely used in applied research studies [26,67–79]. Alternative approaches sometimes used to contrast YPLL by sex are to use sex-specific values of 𝒜 corresponding to at-birth male and female life expectancies (e.g. [80,81]) or the sex-specific remaining life expectancy method as employed in Quast et al. and in other applied research studies (e.g. [82–85]) to reflect known underlying differences in the life expectancy between males and females. However, the gap between the male and female life expectancy changes over time [86], varies substantially across countries [87], and are attributable to a myriad of biological, behavioral, and social factors that, after decades of research, are not fully understood [88]. Hence, out of a sex equity ethos not to necessarily normalize lower life expectancy among males, a sentiment shared in other applications [89], but also to provide a consistent comparison of YPLL by sex [90], we decided to use a constant YPLL upper reference age of 𝒜 = 75 to define YPLL for both males and females—a common practice, as previously discussed. We also obtain conservative 95% interval estimates of age-adjusted mortality rates by sex and age-adjusted male-to-female mortality RR’s in the U.S. and in each state using an analogous MC simulation procedure described in Section 2.7, also for ℬ = 1000 iterations.

All MC simulations were performed using the R version 3.6.0 programming language [91]. The code used in our analysis is available upon reasonable request from the corresponding author.

## 3. Results

### 3.1. Blueprint for Interpretation of Results

Tables S1 and S2 in the Supplementary Materials contain the complete results of our analysis. Table S1 presents the percent population shares by sex, total COVID-19 deaths by sex, percentages of total COVID-19 deaths by sex, conservative 95% interval estimates of total COVID-19-attributable YPLL by sex, and conservative 95% interval estimates of the percentage of total COVID-19-attributable YPLL by sex in the U.S. and in each state. When the percentage of total deaths for males is above their percent population share, males are overrepresented among COVID-19 deaths, and when the percentage of total deaths for males is below their percent population share, males are underrepresented among COVID-19 deaths. The interpretation of the interval estimates of the percentage of total YPLL relative to the percent population share is slightly more nuanced, however. When the interval estimate of the male percentage of total YPLL is completely above the male percent population share, males are either overrepresented among COVID-19 deaths or male decedent ages are systematically younger relative to those of females—to a degree that is statistically discernable—or both. Conversely, when the interval estimate of the male percentage of total YPLL is completely below the male percent population share, males are either underrepresented among COVID-19 deaths or male decedent ages are systematically older relative to those of females—to a degree that is statistically discernable—or both.

The magnitudes of the sex disparities in the COVID-19 mortality burden can be amplified when mortality is measured in terms of YPLL compared to (age-irrespective) death counts. For example, if males are overrepresented among COVID-19 deaths, their interval estimate of the percentage of total YPLL can be completely above their percentage of total deaths as a result of male decedent ages being systematically and statistically discernably younger relative to those of females. Similarly, if males are underrepresented among COVID-19 deaths, their interval estimate of the percentage of total YPLL can be completely below their percentage of total deaths as a result of male decedent ages being systematically and statistically discernably older relative to those of females.

Moreover, the direction of the disparity in the COVID-19 mortality burden can in fact reverse when mortality is measured in terms of YPLL compared to (age-irrespective) death counts. For example, if males are underrepresented among COVID-19 deaths, the interval estimate of the percentage of total YPLL can, in contrast, be completely above the male percent population share as a result of male decedent ages being systematically younger relative to females to a degree that is both statistically discernable and outweighs the disproportionately low number of male deaths. Similarly, if males are overrepresented among COVID-19 deaths, the interval estimate of the percentage of total YPLL can, in contrast, be completely below the male percent population share as a result of male decedent ages being systematically older relative to females to a degree that is both statistically discernable and outweighs the disproportionately high number of male deaths. This second scenario, however, does not occur in the results of our analysis.

Table S2 presents conservative 95% interval estimates of the male and female age-adjusted mortality and YPLL rates as well as the age-adjusted male-to-female mortality and YPLL RR’s in the U.S. and in each state. When the interval estimate of the age-adjusted male-to-female mortality RR is completely above 1.0, it means that after accounting for differences in the male and female population age distributions, the male mortality rate is statistically discernably above that of females. Similarly, when the interval estimate of the age-adjusted male-to-female YPLL RR is completely above 1.0, it means that after accounting for differences in the male and female population age distributions, the male YPLL rate is statistically discernably above that of females. When the interval estimate of the age-adjusted male-to-female YPLL RR is completely above the interval estimate of the age-adjusted male-to-female mortality RR, the male-female disparity in the COVID-19 mortality burden is statistically discernably greater in magnitude when measuring mortality in terms of YPLL compared to (age-irrespective) death counts as a result of males dying at systematically and statistically discernably younger ages relative to females after accounting for differences in their population age distributions.

### 3.2. Presentation of Results

Figure 1 displays a graphical comparison between the conservative 95% interval estimates of the male percentage of total YPLL, the male percentage of total deaths, and the male percent population share in the U.S. and in each state. Nationally, males are overrepresented among COVID-19 deaths, comprising 54.8% of total deaths despite representing 49.2% of the U.S. population. This is also mirrored at the state level, where males are overrepresented among COVID-19 deaths in all but 2 states (Maine and Rhode Island being the exceptions), with percentages of total COVID-19 deaths exceeding the state percent population shares by between 0.03 percentage units in Connecticut to 12.5 percentage units in Nevada. Moreover, males die from COVID-19 in the U.S. overall at systematically younger ages relative to females to such a degree that the U.S. national conservative 95% interval estimate of the male percentage of total YPLL ([64.0–64.1%]) is completely above the U.S. national male percentage of total COVID-19 deaths. This national trend of males dying from COVID-19 at systematically younger ages relative to females is nearly universally observed at the state level, with the interval estimates of the male percentage of total YPLL completely above the male percentage of total deaths in all but 2 states (Hawaii and Alaska being the exceptions). For example, in California, males represent 49.7% of the population and 59.0% of total deaths, but the interval estimate of the percentage of total YPLL is an even higher—and astonishing—[68.3–68.6%]. The direction of the disparity reverses for the two states with males underrepresented among COVID-19 deaths. In Maine, males represent 49.0% of the population and only 48.0% of COVID-19 deaths, but the interval estimate of the percentage of total YPLL is [62.8–68.4%]. Similarly, in Rhode Island, males represent 48.7% of the population and only 48.1% of COVID-19 deaths, but the interval estimate of the percentage of total YPLL is [60.8–64.4%]. Furthermore, in every state except Alaska, the interval estimates of the male percentages of total YPLL are completely above the male percent population shares. To complement Figure 1, Figure 2 displays a graphical comparison between the conservative 95% interval estimates of the female percentage of total YPLL, the female percentage of total deaths, and the female percent population share in the U.S. and in each state.

**Figure 1.**
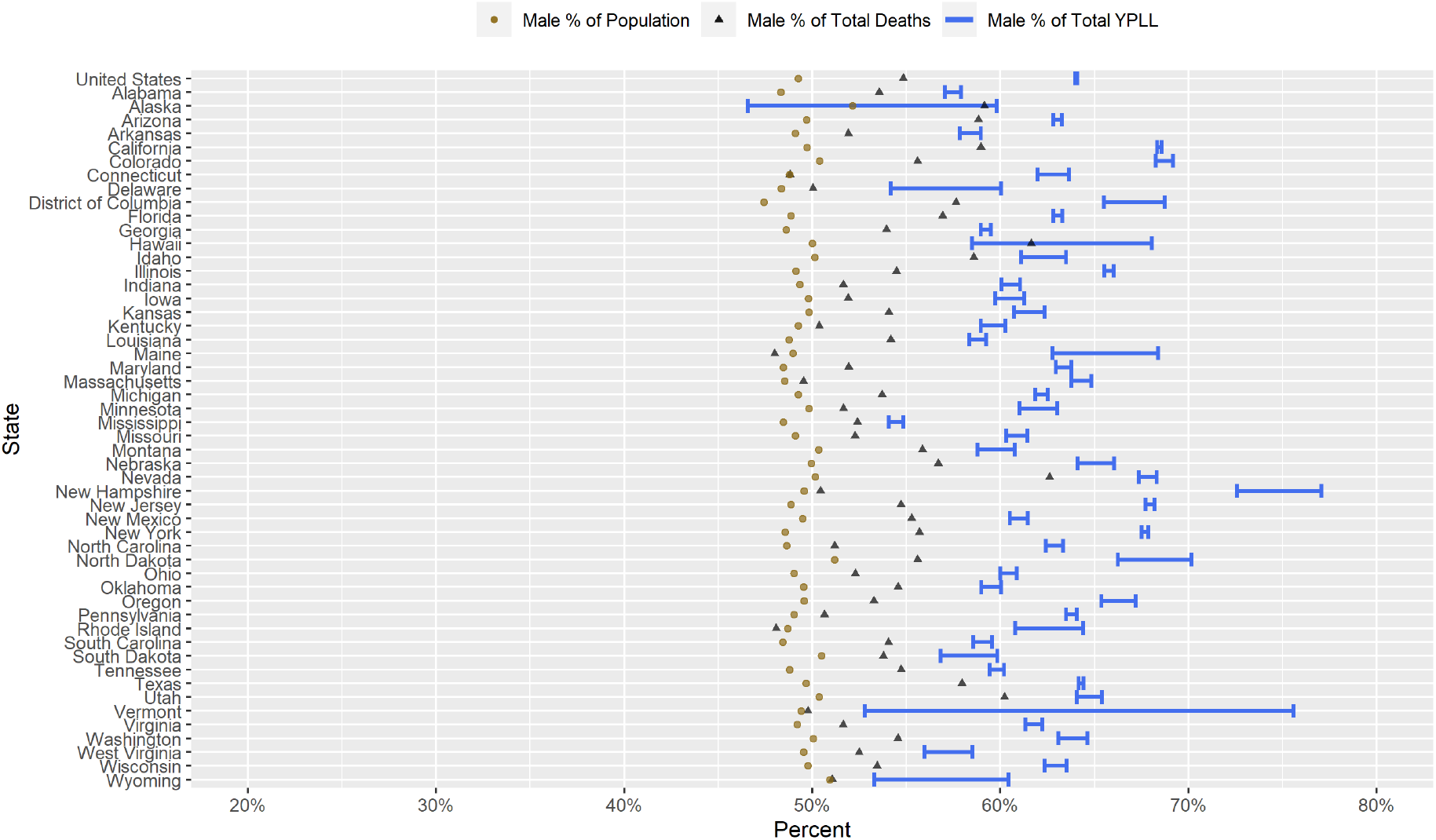
Conservative 95% interval estimates of the percentage of total COVID-19-attributable YPLL before age 75, the percentage of total COVID-19 deaths, and the percent population shares for males in the U.S. and in each of the 50 states and D.C. with respect to cumulative COVID-19 deaths according to data from the National Center for Health Statistics as of 31 March 2021.

**Figure 2.**
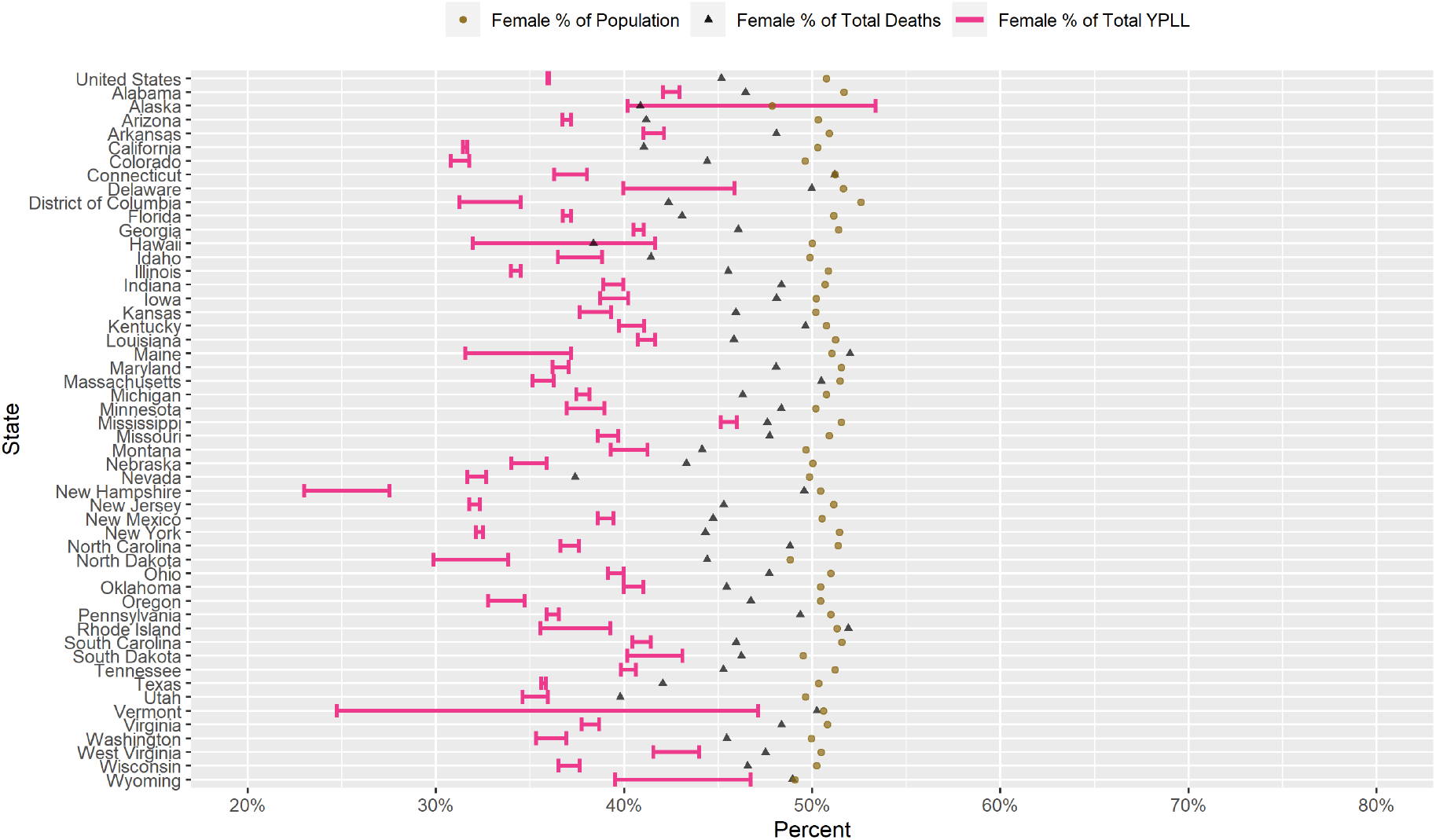
Conservative 95% interval estimates of the percentage of total COVID-19-attributable YPLL before age 75, the percentage of total COVID-19 deaths, and the percent population shares for females in the U.S. and in each of the 50 states and D.C. with respect to cumulative COVID-19 deaths according to data from the National Center for Health Statistics as of 31 March 2021.

Figure 3 presents a graphical comparison of the conservative 95% interval estimates of the age-adjusted male-to-female YPLL and mortality RR’s in the U.S. and in each state. The U.S. national conservative 95% interval estimate of the age-adjusted male-to-female mortality RR is [1.62–1.62], and the state-level interval estimates of the age-adjusted male-to-female mortality RR are completely above 1.0 in every state and completely above 2.0 in Hawaii. Furthermore, after accounting for differences in the male and female national population age distributions, males die from COVID-19 in the U.S. overall at systematically younger ages relative to females to such a degree that the U.S. national conservative 95% interval estimate of the age-adjusted male-to-female YPLL RR ([1.88 – 1.89]) is completely above the U.S. national interval estimate of the male-to-female mortality RR. This national trend is also widely observed at the state level, with the interval estimates of the age-adjusted male-to-female YPLL RR completely above the corresponding interval estimates of the age-adjusted male-to-female mortality RR in 33 states. Intriguingly, the reverse inequality is observed in 4 states (Alabama, Alaska, Mississippi, and South Dakota); hence, for these states, males actually die from COVID-19 at older ages relative to females to a degree that is statistically discernable after accounting for differences in the male and female state population age distributions. For 3 of these 4 states (Alaska being the exception), males actually die from COVID-19 at statistically discernably earlier ages than females without accounting for differences in the male and female state population age distributions (i.e., the interval estimate of the male percentage of total YPLL is completely above the male percentage of total deaths; see Figure 1), thereby illustrating the importance of age-standardization. Nevertheless, for these 3 states, the age-adjusted male YPLL rates still statistically discernably exceed the age-adjusted female YPLL rates due to the disproportionately high number of male COVID-19 deaths in these states. In fact, the state-level interval estimates of the age-adjusted male-to-female YPLL RR are completely above 1.0 in every state except Alaska and, remarkably, are completely above 2.0 in 6 states (California, Colorado, Nevada, New Hampshire, New Jersey, and New York).

**Figure 3.**
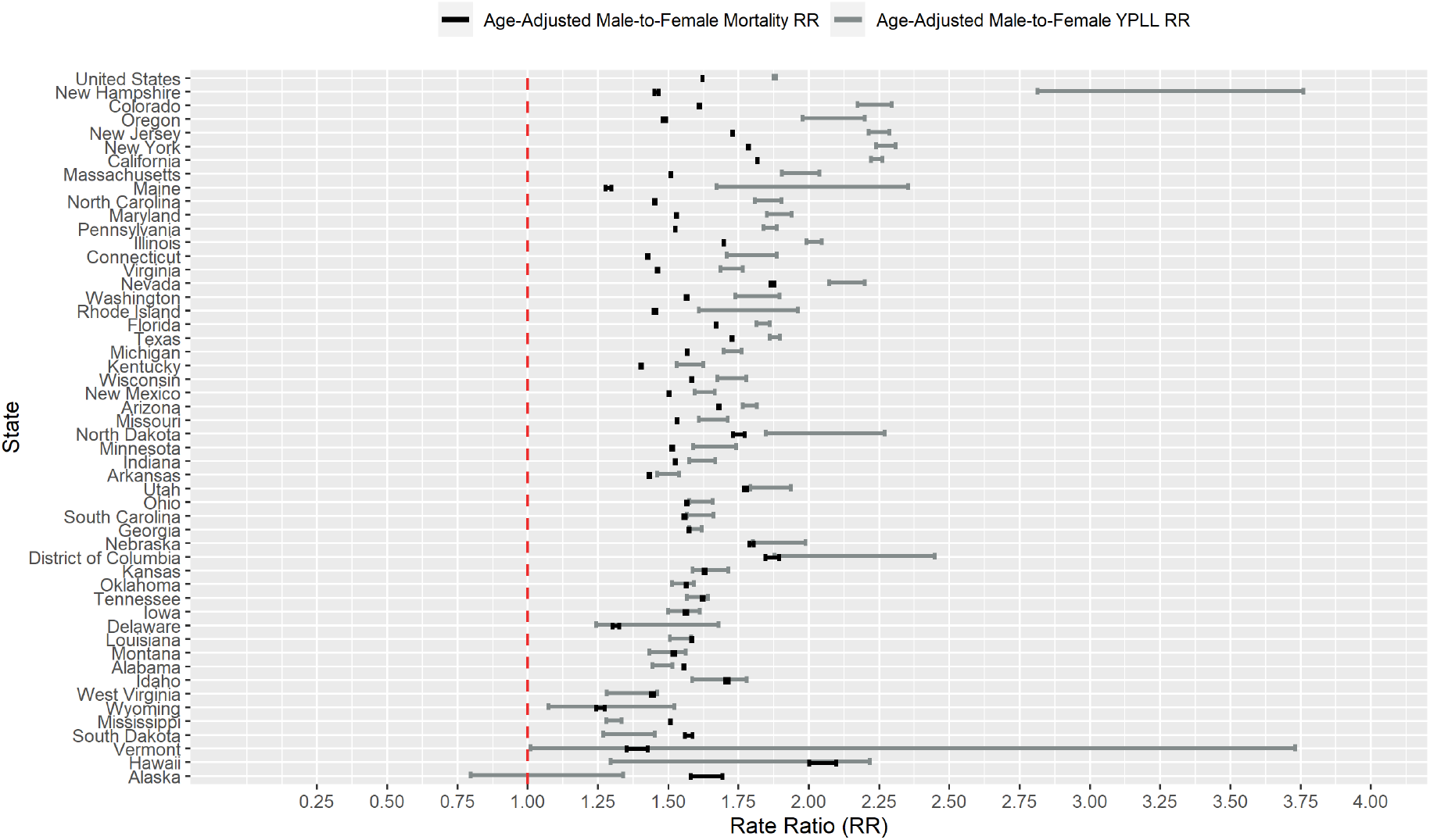
Conservative 95% interval estimates of the age-adjusted male-to-female YPLL and mortality RR’s in the U.S. and in each of the 50 states and D.C. with respect to cumulative COVID-19 deaths according to data from the National Center for Health Statistics as of 31 March 2021. States are ordered from top to bottom in descending order of the signed difference between the lower limit of the YPLL RR interval and the upper limit of the mortality RR interval.

## 4. Discussion

COVID-19 does not affect all segments of the U.S. population equally. As with older individuals [92] and certain racial/ethnic minorities (Blacks, Hispanics, and American Indian and Alaska Natives) [93], men are at disproportionately high risk of hospitalization and mortality following COVID-19 infection [15]. Against this backdrop, it is notable and concerning that U.S. national COVID-19 public health strategies by both the Trump and Biden presidential administrations [94,95] have failed to explicitly acknowledge or propose strategies to address the markedly disproportionate rates of morbidity and mortality associated with COVID-19 infections among men, a trend also observed internationally in governmental pandemic response policies [96].

We echo voices within the public health community calling on policy makers to place greater emphasis on addressing the disproportionate impacts of COVID-19 on men [96–100], especially for the subgroups of men who have the poorest health outcomes, such as certain racial/ethnic minorities. Such efforts should not minimize the need to address serious impacts that COVID-19 has on women, which have been disproportionate in some domains other than morbidity and mortality (e.g. [101–112]), nor should they detract from efforts to improve women’s health more broadly. But the absence of COVID-19 prevention and mitigation efforts focused on men constitutes a glaring missed opportunity to more effectively combat the U.S. COVID-19 epidemic among a substantial proportion of the population. Among the most obvious areas needing attention is the gap in male and female COVID-19 vaccination rates, with notably lower rates among men [113–115]. Vaccine hesitancy among men has emerged as a substantial source of concern, and an NPR/PBS NewsHour/Marist poll conducted 3–8 March 2021 [116] revealed that men were substantially more likely than women to refuse to get a COVID-19 vaccine, with Republican-identifying men especially unwilling (49%). From a public health perspective, the absence of U.S. health policy initiatives specifically targeted to encourage COVID-19 vaccine uptake among men is disconcerting [114,117].

Multiple theories have been proposed attempting to explain the substantial disparities in the COVID-19 mortality burden between males and females, focusing primarily on differences in biological composition, behavior, immunological responses, and comorbidity prevalences between males and females. For example, men have higher rates of certain medical conditions such as coronary artery disease [118,119] and hypertension [120], risk factors for severe illness post-COVID-19 infection [121, 122]. Some research has focused on the mechanisms by which biological differences and differences in immunological responses to COVID-19 infection between males and females contribute to the observed disparities [123,124], with differences in the expressions of angiotensin-converting enzyme-2 (ACE2)—the functional receptor for SARS-CoV-2 [125–127]—and transmembrane serine protease 2 (TMPRSS2)—an enzyme that facilitates SARS-CoV-2 cell entry and spread [127]—by sex, as well as differences in the degree and nature of T-cell activation post-COVID-19 infection between males and females, two particular areas of focus [128–136]. Behavioral differences between men and women have also been proposed [137–139] as contributing to the disparities in the COVID-19 mortality burden by sex. For example, men are more likely to smoke cigarettes [140]—a risk factor for severe illness post-COVID-19 infection [121]—and may seek medical care later in the course of a COVID-19 infection compared to women [141]. Men are also more likely than women to eschew wearing masks [137,142–145], which may reduce the severity of a potential COVID-19 infection by reducing the viral inoculum [146–149]. More broadly, our findings reveal noticeable state-to-state variability in the magnitudes of the estimated disparities, as illustrated in Figures 1-3, which seem to suggest that factors related to social determinants of health [150,151], whose degree of association with sex varies state to state, play a role in driving male-female disparities in COVID-19 mortality, a perspective similarly shared by other COVID-19 researchers [139,152]. For example, men are vastly overrepresented in certain essential industries such as food/agriculture, transportation/logistics, and manufacturing [153], which have been documented as being among the employment domains with the highest associated levels of excess mortality attributable to COVID-19 [154], and these patterns in employment by sex vary by state. While the full scope of factors causing the disproportionately high degree of COVID-19 mortality experienced by males are likely complex, multifaceted, and interactive [97,155], an important limitation of our analysis is that it is a descriptive epidemiological study [156]. We urge further medical research in treatments for COVID-19 infections that can improve outcomes for men as well as recommend more in-depth investigations attempting to pinpoint the social mechanisms that contribute to higher degrees of disparities in some states and lower degrees of disparities in other states, which could inform effective strategies for public health interventions targeting men.

Contrasting the male and female COVID-19 mortality burden using YPLL captures disparities in both the number of COVID-19 deaths and the ages at death of COVID-19 decedents in a single metric that complements conventional comparative COVID-19 mortality analyses by sex, and our results show that measuring mortality in terms of YPLL compared to death counts generally amplifies the magnitude of the disparities in the COVID-19 mortality burden between males and females. For instance, after accounting for the differences in the male and female national population age distributions, we estimated the COVID-19-attributable mortality rate in the U.S. to be approximately 62% higher for males than females but the U.S. national COVID-19-attributable YPLL rate to be 88–89% higher for males than females, owing to the fact that, nationally, males die from COVID-19 at systematically younger ages than females. Remarkably, the age-adjusted male YPLL rates are estimated to be more than twice that of females in 6 states. Furthermore, while substantial disparities in the COVID-19 mortality burden exist between males and females, the overwhelming majority of COVID-19 infected individuals do not die from the disease. However, the long-term health effects of recovered COVID-19 patients are unknown at the time of writing [157], but when more detailed information on the long-term disability profiles of formerly COVID-19-infected individuals becomes available, a comparative analysis of disability-adjusted life years (DALY) [158] by sex, for example, would vastly broaden our understanding of the disparate impacts of the U.S. COVID-19 epidemic between males and females beyond just immediate mortality.

Two sources of uncertainty within the NCHS data substantially complicated our analysis: the administrative interval censoring of ages at death (precluding the exact calculation of YPLL) and the suppression of death counts between 1 and 9 within age groups denoting age at death. To perform estimation of the YPLL-based estimands of interest, accounting for the estimation uncertainty arising from the administrative interval censoring of ages at death and the suppression of low death counts, we developed a novel adaptation of the MC simulation procedure developed by Xu et al. (2021a) that targets estimation of the extrema of the theoretically attainable values of the YPLL-based estimands of interest, resulting in interval estimates for them. A consequence of this conservative estimation strategy, however, was wide interval estimates in states with a high ratio of suppressed to non-suppressed death counts, corresponding to states with a low total number of COVID-19 deaths (e.g., see results for Hawaii). As a result, disparities in COVID-19-attributable YPLL between males and females can be hard to detect when they exist and are small. Despite the challenges in estimation of the YPLL-based estimands of interest due to the two sources of uncertainty in the data and our conservative estimation approach, our analysis nevertheless revealed substantial and statistically discernable disparities in COVID-19-attributable YPLL before age 75 between males and females across U.S. states.

We include a brief methodological discussion here to address our choice of using a constant value of 𝒜 for males and females to define YPLL—a common practice in applied research, as previously discussed. This approach may raise concerns that the estimated male-female disparities in YPLL we claim to be (exclusively) attributable to COVID-19 are in fact partially attributable to non-COVID-19 related factors that account for the underlying male-female life expectancy gap in the overall U.S. population. However, let’s carefully scrutinize the definition of YPLL used in our study. If a COVID-19 death occurs at age *a*_*i*_ that is below 𝒜, that decedent—in principal—had 𝒜 − *a*_*i*_ potential years of life remaining before age 𝒜 if not for premature death by COVID-19. This is true for both male and female decedents even if there is valid reason to suspect the counterfactual number of remaining years of life systematically differs by sex. Hence, our choice of definition of YPLL should be viewed as attempting not to explicitly approximate the counterfactual number of remaining years of life for each decedent that takes into account decedent age and sex, but as calculating a more abstract quantity, namely, the potential—or in other words, theoretically attainable—years of life remaining up to the age of 75 if not for premature death by COVID-19.

As previously discussed in Section 2.2, Quast et al. used the sex-specific remaining life expectancy method to define YPLL. However, the context of COVID-19 reveals an important methodological concern of this approach. The health profiles of COVID-19 decedents are not representative of the overall U.S. population; for example, hospitalized COVID-19 patients have substantially higher rates of obesity [159] and other pre-existing health conditions [160] than the overall population. As such, YPLL estimates using the sex-specific remaining life expectancy method would be expected to vastly overestimate the counterfactual years of life remaining, a point also noted by Quast et al. Furthermore, in light of this, it may not be prudent to readily assume that the counterfactual male-female life expectancy gap among COVID-19 decedents—unascertainable from publicly available data—mirrors that of the overall U.S. population. Indeed, Quast et al. applied a 25% reduction to their estimates of male and female YPLL to reflect this information, but it is unclear why 25% was specifically chosen as the discount factor and why it was the same for males and females. These issues, in addition to concerns previously expressed in Section 2.8 related to sex equity and the desire to provide a consistent comparison of YPLL by sex, constitute the bulk of our resistance to using the sex-specific remaining life expectancy method to define YPLL in this study context.

## 5. Conclusions

We quantified and contrasted COVID-19-attributable YPLL before the age of 75 between males and females in the U.S. and in each of the 50 U.S. states and D.C. from U.S. COVID-19 mortality data from the NCHS (as of 31 March 2021), estimating percentages of total YPLL by sex—contrasting them with their respective percent population shares—and age-adjusted male-to-female YPLL RR’s. Our results revealed a virtually universal pattern across states of males experiencing disproportionately high COVID-19-attributable YPLL relative to females. To examine differences in the characterization of the disparities in the COVID-19 mortality burden between males and females when measuring mortality in terms of YPLL compared to (age-irrespective) death counts, we also calculated the corresponding percentages of total COVID-19 deaths by sex and estimated the corresponding age-adjusted male-to-female mortality RR’s in the U.S. and in each of the 50 states and D.C. Comparing these two approaches to measuring mortality revealed that the estimated disparities are generally greater in magnitude when measuring mortality in terms of YPLL compared to death counts, reflecting a broad dual pattern of males dying from COVID-19 in the U.S. at higher rates and at systematically earlier ages relative to females. As an epidemiological measure, YPLL offers a compelling illustration of the disproportionately high COVID-19 mortality burden experienced by males in the U.S. by explicitly incorporating age at death in quantifying mortality impact.

More broadly, the COVID-19 pandemic offers lessons regarding the importance of cultivating public-health environments in the U.S. and across the world that appropriately recognize the sex-specific needs of individuals as well as different patterns in risk factors, health behaviors, and responses to interventions between men and women. In particular, there is an immediate and urgent need to address COVID-19 vaccine hesitancy among men in the U.S. We urge public officials to update vaccine rollout plans with focused efforts to increase vaccinations among men.

## Data Availability

Data referenced in the manuscript are available in the Supplementary Materials.

## Supplementary Materials

File S1: Cumulative COVID-19 death counts in the U.S. stratified by state, sex, and age group from the National Center for Health Statistics as of 31 March 2021, File S2: 2019 U.S. Census Bureau estimates of the U.S. population stratified by state, sex, and age, Table S1: Percent population shares by sex, total COVID-19 deaths by sex, percentages of total COVID-19 deaths by sex, conservative 95% interval estimates of total COVID-19-attributable YPLL before age 75 by sex, and conservative 95% interval estimates of the percentage of total COVID-19-attributable YPLL before age 75 by sex in the U.S. and in each state and D.C., Table S2: Conservative 95% interval estimates of age-adjusted mortality and YPLL rates by sex and conservative 95% interval estimates of age-adjusted male-to-female mortality and YPLL rate ratios in the U.S. and in each state and D.C.

## Author Contributions

Conceptualization, J.J.X.; Data curation, J.J.X.; Formal analysis, J.J.X.; Methodology, J.J.X.; Software, J.J.X.; Visualization, J.J.X.; Writing – original draft, J.J.X.; Writing – review & editing, J.J.X., J.T.C., T.R.B., R.S.B., M.A.S. and C.M.R.

## Abbreviations

The following abbreviations are used in this manuscript:

CDC: Centers for Disease Control and Prevention
COVID-19: Coronavirus Disease 2019
D.C.: District of Columbia
MC: Monte Carlo
NCHS: National Center for Health Statistics
RR: Rate Ratio
SARS-CoV-2: Severe Acute Respiratory Syndrome Coronavirus 2
U.S.: United States
YPLL: Years of Potential Life Lost

