## Supplementary material for "Male-Female Disparities in Years of Potential Life Lost Attributable to COVID-19 in the United States: A State-by-State Analysis": Table S1

| State | % Population | | Total Deaths | | % Deaths | | Total YPLL | | % YPLL | |
|---|---|---|---|---|---|---|---|---|---|---|
| | Male | Female | Male | Female | Male | Female | Male | Female | Male | Female |
| US | 49.2% | 50.8% | 292,402 | 240,889 | 54.8% | 45.2% | [1,741,317 – 1,745,559] | [977,972 – 981,179] | [64.0% – 64.1%] | [35.9% – 36.0%] |
| AL | 48.3% | 51.7% | 5,383 | 4,669 | 53.6% | 46.4% | [33,004 – 33,727] | [24,366 – 24,977] | [57.0% – 57.9%] | [42.1% – 42.9%] |
| AK | 52.1% | 47.9% | 181 | 125 | 59.2% | 40.8% | [1,058 – 1,321] | [867 – 1,244] | [46.6% – 59.8%] | [40.2% – 53.4%] |
| AZ | 49.7% | 50.3% | 8,925 | 6,245 | 58.8% | 41.2% | [62,384 – 63,134] | [36,474 – 37,086] | [62.8% – 63.3%] | [36.7% – 37.2%] |
| AR | 49.1% | 50.9% | 2,985 | 2,765 | 51.9% | 48.1% | [15,927 – 16,322] | [11,275 – 11,687] | [57.8% – 59.0%] | [41.0% – 42.1%] |
| CA | 49.7% | 50.3% | 35,500 | 24,710 | 59.0% | 41.0% | [274,859 – 276,443] | [126,535 – 127,684] | [68.3% – 68.6%] | [31.4% – 31.7%] |
| CO | 50.4% | 49.6% | 3,468 | 2,770 | 55.6% | 44.4% | [18,793 – 19,282] | [8,516 – 8,815] | [68.2% – 69.2%] | [30.8% – 31.8%] |
| CT | 48.8% | 51.2% | 3,729 | 3,909 | 48.8% | 51.2% | [15,695 – 16,127] | [9,110 – 9,711] | [62.0% – 63.7%] | [36.3% – 38.0%] |
| DE | 48.3% | 51.7% | 699 | 698 | 50.0% | 50.0% | [3,443 – 4,033] | [2,637 – 2,963] | [54.2% – 60.1%] | [40.0% – 45.9%] |
| DC | 47.4% | 52.6% | 762 | 560 | 57.6% | 42.4% | [7,264 – 7,808] | [3,517 – 3,862] | [65.5% – 68.7%] | [31.2% – 34.5%] |
| FL | 48.9% | 51.1% | 17,585 | 13,304 | 56.9% | 43.1% | [90,952 – 91,920] | [53,172 – 53,988] | [62.8% – 63.3%] | [36.7% – 37.2%] |
| GA | 48.6% | 51.4% | 8,706 | 7,434 | 53.9% | 46.1% | [56,651 – 57,451] | [38,975 – 39,636] | [59.0% – 59.5%] | [40.5% – 41.0%] |
| HI | 50.0% | 50.0% | 270 | 168 | 61.6% | 38.4% | [1,985 – 2,287] | [1,046 – 1,440] | [58.5% – 68.0%] | [31.9% – 41.7%] |
| ID | 50.1% | 49.9% | 1,143 | 808 | 58.6% | 41.4% | [4,332 – 4,630] | [2,623 – 2,796] | [61.1% – 63.5%] | [36.5% – 38.8%] |
| IL | 49.1% | 50.9% | 11,124 | 9,298 | 54.5% | 45.5% | [64,555 – 65,350] | [33,506 – 34,120] | [65.5% – 66.0%] | [34.0% – 34.5%] |
| IN | 49.3% | 50.7% | 6,144 | 5,751 | 51.7% | 48.3% | [26,284 – 26,929] | [17,033 – 17,567] | [60.1% – 61.0%] | [38.9% – 40.0%] |
| IA | 49.8% | 50.2% | 3,020 | 2,798 | 51.9% | 48.1% | [11,319 – 11,683] | [7,306 – 7,692] | [59.7% – 61.3%] | [38.7% – 40.2%] |
| KA | 49.8% | 50.2% | 2,563 | 2,177 | 54.1% | 45.9% | [11,850 – 12,205] | [7,309 – 7,734] | [60.7% – 62.4%] | [37.6% – 39.3%] |
| KY | 49.3% | 50.7% | 3,393 | 3,343 | 50.4% | 49.6% | [16,108 – 16,752] | [10,944 – 11,300] | [59.0% – 60.3%] | [39.7% – 41.1%] |
| LA | 48.8% | 51.2% | 4,868 | 4,119 | 54.2% | 45.8% | [30,379 – 30,969] | [21,180 – 21,801] | [58.4% – 59.2%] | [40.7% – 41.6%] |
| ME | 49.0% | 51.0% | 396 | 429 | 48.0% | 52.0% | [1,467 – 1,658] | [740 – 899] | [62.8% – 68.4%] | [31.6% – 37.2%] |
| MD | 48.4% | 51.6% | 4,775 | 4,421 | 51.9% | 48.1% | [28,946 – 29,632] | [16,700 – 17,159] | [63.0% – 63.8%] | [36.2% – 37.1%] |
| MA | 48.5% | 51.5% | 6,349 | 6,471 | 49.5% | 50.5% | [23,533 – 24,202] | [13,030 – 13,472] | [63.8% – 64.8%] | [35.2% – 36.3%] |
| MI | 49.3% | 50.7% | 7,966 | 6,865 | 53.7% | 46.3% | [41,166 – 41,820] | [24,930 – 25,525] | [61.8% – 62.5%] | [37.5% – 38.1%] |
| MN | 49.8% | 50.2% | 3,634 | 3,401 | 51.7% | 48.3% | [13,054 – 13,433] | [7,805 – 8,388] | [61.0% – 63.0%] | [37.0% – 39.0%] |
| MS | 48.5% | 51.5% | 3,701 | 3,363 | 52.4% | 47.6% | [23,223 – 23,788] | [19,461 – 19,875] | [54.1% – 54.8%] | [45.1% – 46.0%] |
| MO | 49.1% | 50.9% | 5,416 | 4,945 | 52.3% | 47.7% | [24,381 – 24,991] | [15,590 – 16,161] | [60.3% – 61.4%] | [38.6% – 39.7%] |
| MT | 50.3% | 49.7% | 863 | 682 | 55.9% | 44.1% | [3,999 – 4,211] | [2,679 – 2,849] | [58.8% – 60.8%] | [39.3% – 41.3%] |
| NE | 50.0% | 50.0% | 1,557 | 1,189 | 56.7% | 43.3% | [7,253 – 7,553] | [3,836 – 4,112] | [64.1% – 66.0%] | [34.0% – 35.9%] |
| NV | 50.2% | 49.8% | 3,332 | 1,990 | 62.6% | 37.4% | [23,858 – 24,417] | [11,227 – 11,652] | [67.3% – 68.3%] | [31.7% – 32.7%] |
| NH | 49.6% | 50.4% | 628 | 617 | 50.4% | 49.6% | [1,857 – 2,137] | [617 – 726] | [72.6% – 77.1%] | [23.0% – 27.5%] |
| NJ | 48.9% | 51.1% | 12,297 | 10,181 | 54.7% | 45.3% | [79,426 – 80,326] | [37,272 – 38,091] | [67.7% – 68.2%] | [31.8% – 32.3%] |
| NM | 49.5% | 50.5% | 2,079 | 1,682 | 55.3% | 44.7% | [17,228 – 17,648] | [10,995 – 11,307] | [60.5% – 61.5%] | [38.6% – 39.5%] |
| NY | 48.6% | 51.4% | 28,115 | 22,372 | 55.7% | 44.3% | [181,560 – 182,990] | [86,418 – 87,571] | [67.5% – 67.9%] | [32.1% – 32.5%] |
| NC | 48.6% | 51.4% | 3,265 | 3,113 | 51.2% | 48.8% | [19,564 – 20,095] | [11,528 – 11,885] | [62.4% – 63.3%] | [36.6% – 37.6%] |
| ND | 51.2% | 48.8% | 909 | 726 | 55.6% | 44.4% | [3,971 – 4,240] | [1,780 – 2,060] | [66.3% – 70.2%] | [29.9% – 33.8%] |
| OH | 49.0% | 51.0% | 10,895 | 9,943 | 52.3% | 47.7% | [43,367 – 44,179] | [28,293 – 29,059] | [60.0% – 60.9%] | [39.1% – 40.0%] |
| OK | 49.5% | 50.5% | 4,295 | 3,578 | 54.6% | 45.4% | [23,875 – 24,514] | [16,204 – 16,690] | [59.0% – 60.0%] | [40.0% – 41.0%] |
| OR | 49.6% | 50.4% | 1,173 | 1,029 | 53.3% | 46.7% | [6,013 – 6,319] | [3,041 – 3,237] | [65.4% – 67.2%] | [32.8% – 34.7%] |
| PA | 49.0% | 51.0% | 12,964 | 12,635 | 50.6% | 49.4% | [54,222 – 54,981] | [30,703 – 31,304] | [63.5% – 64.1%] | [35.9% – 36.5%] |
| RI | 48.7% | 51.3% | 1,216 | 1,313 | 48.1% | 51.9% | [4,696 – 4,933] | [2,677 – 3,083] | [60.8% – 64.4%] | [35.6% – 39.3%] |
| SC | 48.4% | 51.6% | 4,397 | 3,738 | 54.1% | 45.9% | [23,763 – 24,441] | [16,483 – 16,943] | [58.5% – 59.5%] | [40.4% – 41.4%] |
| SD | 50.5% | 49.5% | 1,068 | 918 | 53.8% | 46.2% | [4,567 – 4,781] | [3,165 – 3,518] | [56.8% – 59.8%] | [40.2% – 43.1%] |
| TN | 48.8% | 51.2% | 6,481 | 5,362 | 54.7% | 45.3% | [36,766 – 37,538] | [24,687 – 25,278] | [59.4% – 60.2%] | [39.8% – 40.6%] |
| TX | 49.7% | 50.3% | 29,171 | 21,163 | 58.0% | 42.0% | [225,693 – 227,235] | [125,188 – 126,369] | [64.2% – 64.4%] | [35.6% – 35.8%] |
| UT | 50.4% | 49.6% | 1,359 | 898 | 60.2% | 39.8% | [8,876 – 9,236] | [4,828 – 5,044] | [64.1% – 65.4%] | [34.6% – 35.9%] |
| VT | 49.4% | 50.6% | 104 | 105 | 49.8% | 50.2% | [354 – 577] | [181 – 330] | [52.8% – 75.6%] | [24.7% – 47.1%] |
| VA | 49.2% | 50.8% | 5,102 | 4,778 | 51.6% | 48.4% | [25,855 – 26,491] | [15,985 – 16,419] | [61.3% – 62.2%] | [37.7% – 38.7%] |
| WA | 50.1% | 49.9% | 2,658 | 2,215 | 54.5% | 45.5% | [13,484 – 13,872] | [7,512 – 7,966] | [63.1% – 64.6%] | [35.3% – 36.9%] |
| WV | 49.5% | 50.5% | 1,320 | 1,195 | 52.5% | 47.5% | [5,347 – 5,761] | [4,038 – 4,264] | [56.0% – 58.5%] | [41.5% – 44.0%] |
| WI | 49.8% | 50.2% | 4,155 | 3,620 | 53.4% | 46.6% | [16,725 – 17,257] | [9,840 – 10,170] | [62.4% – 63.5%] | [36.5% – 37.6%] |
| WY | 50.9% | 49.1% | 314 | 301 | 51.1% | 48.9% | [1,527 – 1,734] | [1,107 – 1,373] | [53.3% – 60.4%] | [39.5% – 46.7%] |

**Table S1**: Percentages of total population, total COVID-19 deaths, percentages of total COVID-19 deaths, conservative 95% interval estimates of total YPLL, and conservative 95% interval estimates of the percentage of total YPLL by sex in the U.S. and in each state and D.C. with respect to cumulative COVID-19 deaths according to data from the National Center for Health Statistics as of 31 March 2021. The upper reference age used to define YPLL is 75 years.
