## Supplementary material for "Male-Female Disparities in Years of Potential Life Lost Attributable to COVID-19 in the United States: A State-by-State Analysis": Table S2

| State | Mortality Rate (per 10,000) | | | YPLL Rate (per 10,000) | | |
|---|---|---|---|---|---|---|
| | Male | Female | Ratio | Male | Female | Ratio |
| US | [20.7 – 20.7] | [12.7 – 12.7] | [1.62 – 1.62] | [117.1 – 117.4] | [62.3 – 62.5] | [1.87 – 1.88] |
| AL | [25.2 – 25.2] | [16.2 – 16.2] | [1.55 – 1.56] | [148.4 – 152.3] | [100.1 – 103.1] | [1.44 – 1.51] |
| AK | [7.8 – 8.1] | [4.9 – 4.9] | [1.58 – 1.69] | [30.6 – 36.7] | [27.0 – 39.1] | [0.80 – 1.34] |
| AZ | [25.6 – 25.6] | [15.2 – 15.2] | [1.68 – 1.68] | [194.7 – 197.9] | [108.8 – 110.6] | [1.77 – 1.81] |
| AR | [22.3 – 22.3] | [15.6 – 15.6] | [1.43 – 1.43] | [118.5 – 121.4] | [78.5 – 81.7] | [1.46 – 1.54] |
| CA | [21.9 – 21.9] | [12.0 – 12.0] | [1.82 – 1.82] | [155.8 – 157.1] | [69.4 – 70.2] | [2.22 – 2.26] |
| CO | [16.2 – 16.2] | [10.0 – 10.1] | [1.61 – 1.61] | [71.5 – 73.7] | [31.9 – 33.1] | [2.17 – 2.30] |
| CT | [22.5 – 22.5] | [15.8 – 15.8] | [1.42 – 1.43] | [92.8 – 95.5] | [50.2 – 54.8] | [1.71 – 1.89] |
| DE | [13.9 – 13.9] | [10.6 – 10.6] | [1.30 – 1.32] | [75.6 – 90.1] | [53.1 – 61.4] | [1.24 – 1.68] |
| DC | [32.5 – 32.8] | [17.4 – 17.6] | [1.85 – 1.89] | [262.9 – 289.3] | [119.0 – 140.6] | [1.88 – 2.45] |
| FL | [14.7 – 14.7] | [8.8 – 8.8] | [1.67 – 1.67] | [91.1 – 92.1] | [49.4 – 50.4] | [1.81 – 1.86] |
| GA | [22.7 – 22.7] | [14.4 – 14.4] | [1.57 – 1.58] | [122.6 – 124.9] | [76.9 – 78.1] | [1.57 – 1.62] |
| HI | [3.8 – 3.8] | [1.9 – 1.9] | [2.00 – 2.10] | [30.1 – 35.8] | [16.0 – 23.6] | [1.29 – 2.22] |
| ID | [15.3 – 15.4] | [9.0 – 9.0] | [1.70 – 1.72] | [54.3 – 58.3] | [32.4 – 34.6] | [1.59 – 1.78] |
| IL | [20.9 – 20.9] | [12.3 – 12.3] | [1.70 – 1.70] | [111.6 – 113.4] | [55.3 – 56.3] | [1.99 – 2.05] |
| IN | [22.6 – 22.6] | [14.8 – 14.8] | [1.52 – 1.53] | [86.6 – 89.1] | [53.3 – 55.3] | [1.57 – 1.67] |
| IA | [20.8 – 20.9] | [13.4 – 13.4] | [1.56 – 1.57] | [78.7 – 81.2] | [50.0 – 53.0] | [1.50 – 1.61] |
| KA | [20.4 – 20.4] | [12.5 – 12.5] | [1.63 – 1.63] | [91.7 – 94.4] | [54.6 – 58.2] | [1.59 – 1.71] |
| KY | [18.3 – 18.3] | [13.0 – 13.0] | [1.40 – 1.41] | [77.9 – 81.3] | [49.6 – 51.3] | [1.53 – 1.62] |
| LA | [25.8 – 25.8] | [16.3 – 16.3] | [1.58 – 1.59] | [147.0 – 150.7] | [95.0 – 98.2] | [1.50 – 1.58] |
| ME | [5.6 – 5.6] | [4.4 – 4.4] | [1.28 – 1.30] | [21.3 – 24.4] | [10.1 – 13.0] | [1.67 – 2.35] |
| MD | [19.4 – 19.4] | [12.7 – 12.7] | [1.53 – 1.53] | [104.9 – 108.0] | [55.5 – 56.9] | [1.85 – 1.94] |
| MA | [21.5 – 21.6] | [14.3 – 14.3] | [1.51 – 1.51] | [73.9 – 76.6] | [37.5 – 39.1] | [1.90 – 2.04] |
| MI | [17.6 – 17.6] | [11.2 – 11.2] | [1.57 – 1.57] | [87.3 – 88.7] | [50.2 – 51.7] | [1.70 – 1.76] |
| MN | [14.7 – 14.7] | [9.7 – 9.7] | [1.51 – 1.52] | [50.1 – 51.5] | [29.4 – 31.7] | [1.59 – 1.74] |
| MS | [30.5 – 30.5] | [20.2 – 20.2] | [1.51 – 1.51] | [176.5 – 181.8] | [135.7 – 138.8] | [1.28 – 1.33] |
| MO | [19.9 – 19.9] | [13.0 – 13.0] | [1.53 – 1.53] | [86.6 – 89.3] | [51.9 – 54.1] | [1.61 – 1.71] |
| MT | [16.1 – 16.1] | [10.6 – 10.6] | [1.51 – 1.52] | [78.2 – 82.3] | [51.9 – 55.4] | [1.43 – 1.56] |
| NE | [18.7 – 18.8] | [10.4 – 10.5] | [1.79 – 1.80] | [84.6 – 88.5] | [44.2 – 47.4] | [1.80 – 1.99] |
| NV | [24.8 – 24.8] | [13.2 – 13.3] | [1.86 – 1.88] | [166.2 – 170.9] | [77.3 – 80.5] | [2.07 – 2.20] |
| NH | [10.1 – 10.1] | [6.9 – 6.9] | [1.45 – 1.47] | [26.7 – 30.9] | [8.2 – 9.6] | [2.81 – 3.76] |
| NJ | [31.4 – 31.5] | [18.3 – 18.3] | [1.73 – 1.73] | [191.5 – 193.7] | [84.4 – 86.8] | [2.21 – 2.29] |
| NM | [21.2 – 21.2] | [14.1 – 14.1] | [1.50 – 1.51] | [183.9 – 188.9] | [112.7 – 115.8] | [1.59 – 1.67] |
| NY | [32.5 – 32.5] | [18.2 – 18.2] | [1.78 – 1.79] | [204.9 – 207.4] | [90.3 – 92.0] | [2.24 – 2.31] |
| NC | [7.5 – 7.6] | [5.2 – 5.2] | [1.45 – 1.45] | [41.2 – 42.5] | [22.2 – 22.9] | [1.81 – 1.90] |
| ND | [27.5 – 27.5] | [15.8 – 15.8] | [1.73 – 1.77] | [116.8 – 126.7] | [55.1 – 63.9] | [1.85 – 2.27] |
| OH | [21.1 – 21.1] | [13.5 – 13.5] | [1.56 – 1.57] | [79.4 – 81.3] | [48.9 – 50.7] | [1.58 – 1.66] |
| OK | [25.9 – 25.9] | [16.5 – 16.6] | [1.56 – 1.57] | [138.3 – 142.4] | [89.1 – 92.0] | [1.51 – 1.59] |
| OR | [6.2 – 6.2] | [4.1 – 4.2] | [1.48 – 1.49] | [30.6 – 32.6] | [14.7 – 15.6] | [1.98 – 2.20] |
| PA | [21.1 – 21.1] | [13.8 – 13.9] | [1.52 – 1.53] | [89.0 – 90.3] | [47.6 – 48.6] | [1.84 – 1.89] |
| RI | [25.5 – 25.5] | [17.6 – 17.6] | [1.45 – 1.46] | [94.8 – 99.8] | [50.0 – 60.0] | [1.61 – 1.96] |
| SC | [19.3 – 19.4] | [12.4 – 12.4] | [1.55 – 1.56] | [101.0 – 104.5] | [62.7 – 64.8] | [1.57 – 1.66] |
| SD | [26.9 – 27.0] | [17.1 – 17.2] | [1.56 – 1.59] | [111.5 – 116.9] | [79.4 – 88.9] | [1.27 – 1.45] |
| TN | [22.6 – 22.6] | [13.9 – 13.9] | [1.62 – 1.63] | [118.2 – 121.2] | [73.6 – 75.8] | [1.57 – 1.64] |
| TX | [28.4 – 28.5] | [16.5 – 16.5] | [1.73 – 1.73] | [184.1 – 185.8] | [97.8 – 99.1] | [1.86 – 1.90] |
| UT | [13.4 – 13.4] | [7.5 – 7.6] | [1.77 – 1.78] | [71.9 – 75.2] | [38.7 – 40.4] | [1.79 – 1.94] |
| VT | [3.5 – 3.6] | [2.5 – 2.6] | [1.35 – 1.43] | [11.1 – 20.7] | [5.4 – 11.5] | [1.01 – 3.73] |
| VA | [14.8 – 14.8] | [10.1 – 10.1] | [1.46 – 1.46] | [66.6 – 68.5] | [38.6 – 39.7] | [1.69 – 1.77] |
| WA | [8.7 – 8.7] | [5.5 – 5.5] | [1.56 – 1.57] | [38.7 – 39.8] | [20.8 – 22.4] | [1.74 – 1.90] |
| WV | [14.3 – 14.4] | [9.9 – 10.0] | [1.44 – 1.45] | [58.7 – 64.4] | [43.9 – 46.5] | [1.28 – 1.46] |
| WI | [15.8 – 15.9] | [10.0 – 10.0] | [1.58 – 1.59] | [59.7 – 62.1] | [34.8 – 36.0] | [1.67 – 1.78] |
| WY | [12.2 – 12.3] | [9.7 – 9.7] | [1.24 – 1.27] | [55.0 – 63.8] | [41.2 – 52.3] | [1.07 – 1.52] |

**Table S2**: Conservative 95% interval estimates of age-adjusted mortality and YPLL rates by sex and age-adjusted male-to-female mortality and YPLL rate ratios in the U.S. and in each state and D.C. with respect to cumulative COVID-19 deaths according to data from the National Center for Health Statistics as of 31 March 2021. The upper reference age used to define YPLL is 75 years.
